# Therapy Without Borders: A Systematic Review on Telehealth’s Role in Expanding Mental Health Access

**DOI:** 10.1101/2024.07.30.24311208

**Authors:** Jennifer Swint, Margaret Fischer, Wei Zhang, Xi Zhang

## Abstract

**Background:** The COVID-19 pandemic has accelerated the adoption of telehealth services in mental healthcare. This systematic review aims to evaluate the effectiveness of telehealth interventions for mental health conditions compared to traditional face-to-face treatment.

**Methods:** We searched major electronic databases (PubMed, PsycINFO, CINAHL, and Cochrane Library) for randomized controlled trials published between 2010 and 2023. Studies comparing telehealth interventions to face-to-face treatment for adults with mental health disorders were included. Two independent reviewers assessed study quality and extracted data. Meta-analyses were conducted where appropriate.

**Results:** Thirty-five studies met the inclusion criteria, encompassing 4,827 participants across various mental health conditions. Telehealth interventions demonstrated non-inferiority to face-to-face treatment for depression (standardized mean difference [SMD] = -0.03, 95% CI [-0.15, 0.09]) and anxiety disorders (SMD = -0.06, 95% CI [-0.19, 0.07]). For post-traumatic stress disorder, telehealth showed a small but significant advantage (SMD = -0.21, 95% CI [-0.37, -0.05]). Patient satisfaction and therapeutic alliance were comparable between telehealth and face-to-face interventions. However, dropout rates were slightly higher in telehealth conditions (risk ratio = 1.27, 95% CI [1.11, 1.46]).

**Conclusion:** This review suggests that telehealth interventions are generally as effective as face-to-face treatment for common mental health disorders. While promising, these findings should be interpreted cautiously due to heterogeneity in study designs and interventions. Future research should focus on long-term outcomes, cost-effectiveness, and strategies to improve engagement in telehealth settings.

## Introduction

Mental health disorders represent a significant global health challenge, affecting an estimated 970 million people worldwide and accounting for 10.8% of global disability-adjusted life years (DALYs) [1]. Despite the availability of effective treatments, a substantial treatment gap persists, with only a minority of individuals receiving adequate care [2]. Barriers to accessing mental health services are multifaceted, including geographical constraints, stigma, financial limitations, and shortages of trained professionals, particularly in low- and middle-income countries [3, 4].

Telehealth, defined as the use of information and communication technologies to deliver health services at a distance, has emerged as a promising solution to address these challenges in mental healthcare [5]. The modalities of telehealth in mental health, often referred to as telemental health or telepsychiatry, encompass a wide range of interventions, including video conferencing, telephone counseling, web-based interventions, mobile applications, and text messaging [6, 7].

The COVID-19 pandemic has catalyzed an unprecedented acceleration in the adoption of telehealth across healthcare systems globally, with mental health services at the forefront of this digital transformation [8]. This rapid shift has not only demonstrated the feasibility of large-scale telemental health implementation but also highlighted its potential to maintain continuity of care during crises and expand access to mental health services in general [9, 10].

The application of telehealth in mental health spans a broad spectrum of conditions, including mood disorders, anxiety disorders, post-traumatic stress disorder (PTSD), substance use disorders, and severe mental illnesses such as schizophrenia [11, 12]. Early research has shown promising results for telehealth interventions across various mental health conditions, with some studies suggesting comparable efficacy to face-to-face treatments [13, 14].

However, the rapid proliferation of telemental health services has also raised important questions about its effectiveness, acceptability, and implementation challenges. Concerns have been voiced regarding the quality of therapeutic alliance in remote settings, the digital divide exacerbating health inequities, and the potential limitations in assessing and managing acute psychiatric crises [15, 16].

While previous systematic reviews and meta-analyses have examined the efficacy of specific telehealth interventions for particular mental health conditions [17, 18, 19], there is a pressing need for a comprehensive, up-to-date systematic review that evaluates the overall effectiveness of telehealth across various mental health disorders and directly compares it to traditional face-to-face treatment. Such a review is crucial in the context of the post-pandemic era, where hybrid models of care are likely to become the norm [20].

This systematic review aims to address this gap by synthesizing the current evidence on the effectiveness of telehealth interventions for mental health conditions compared to face-to-face treatment. Specifically, we will focus on:

- Clinical outcomes across different mental health disorders
- Patient satisfaction and acceptability
- Therapeutic alliance in remote versus in-person settings
- Engagement and adherence to treatment
- Cost-effectiveness and resource utilization

By providing a comprehensive analysis of the current state of evidence, this review seeks to inform clinical practice, guide policy decisions, and identify areas for future research in the rapidly evolving field of telemental health. Additionally, we aim to explore potential moderators of treatment effectiveness, such as patient characteristics, technology platforms, and specific intervention components, to guide the development of more targeted and effective telemental health interventions.

## Methods

### Search Strategy

We conducted a comprehensive search of four major electronic databases: PubMed, PsycINFO, CINAHL, and Cochrane Library. The search covered studies published between January 1, 2010, and December 31, 2023. We used the following search terms in various combinations:

(“telehealth” OR “telemedicine” OR “telepsychiatry” OR “telemental health” OR “video conferencing” OR “internet-based” OR “web-based” OR “mobile health” OR “mHealth”)

AND

(“mental health” OR “psychiatry” OR “psychology” OR “depression” OR “anxiety” OR “PTSD” OR “substance use” OR “schizophrenia”)

AND

(“randomized controlled trial” OR “RCT” OR “controlled clinical trial”)

Additionally, we hand-searched reference lists of included studies and relevant systematic reviews to identify any missed eligible studies.

## Inclusion and Exclusion Criteria

### Inclusion criteria

1. Randomized controlled trials (RCTs)
2. Studies comparing telehealth interventions to face-to-face treatment
3. Adult participants (≥18 years) with diagnosed mental health disorders
4. Studies published in English
5. Peer-reviewed articles
6. Studies reporting at least one of the following outcomes: symptom severity, quality of life, treatment adherence, patient satisfaction, or cost-effectiveness

### Exclusion criteria

1. Non-randomized studies or observational studies
2. Studies focusing solely on telehealth without a face-to-face comparison group
3. Studies on children or adolescents (<18 years)
4. Studies not reporting primary data (e.g., reviews, meta-analyses)
5. Conference abstracts or unpublished studies
6. Studies with insufficient data for effect size calculation

### Study Selection

Two independent reviewers screened titles and abstracts of identified studies using Covidence systematic review software. Full-text articles of potentially eligible studies were then assessed against the inclusion and exclusion criteria. Disagreements were resolved through discussion with a third reviewer. We calculated Cohen’s kappa to assess inter-rater reliability for the screening process.

### Data Extraction

A standardized data extraction form was developed and piloted on a random sample of 5 included studies. Two reviewers independently extracted data from all included studies. The following information was collected:

1. Study characteristics (author, year, country, study design)
2. Participant demographics (age, gender, ethnicity, mental health condition, severity)
3. Intervention details (type of telehealth, platform used, duration, frequency, provider characteristics)
4. Comparison treatment details (type, duration, frequency)
5. Outcome measures (primary and secondary)
6. Results (including effect sizes, confidence intervals, p-values)
7. Quality assessment indicators
8. Funding source and potential conflicts of interest

### Quality Assessment

The quality of included studies was assessed using the revised Cochrane Risk of Bias tool for randomized trials (RoB 2). This tool evaluates bias across five domains: randomization process, deviations from intended interventions, missing outcome data, measurement of the outcome, and selection of the reported result. Two reviewers independently evaluated each study, with disagreements resolved through discussion or consultation with a third reviewer. We used the GRADE approach to assess the overall quality of evidence for each main outcome.

### Data Synthesis and Analysis

We conducted meta-analyses using a random-effects model for outcomes with sufficient homogeneity across studies. Standardized mean differences (SMD) were calculated for continuous outcomes, and risk ratios (RR) for dichotomous outcomes. 95% confidence intervals were reported for all effect sizes. Heterogeneity was assessed using the I^2^ statistic, with I^2^ values of 25%, 50%, and 75% suggesting low, moderate, and high heterogeneity, respectively. We used the Comprehensive Meta-Analysis software (version 3) for all analyses.

### Subgroup analyses were performed to explore potential sources of heterogeneity, including

1. Type of mental health condition
2. Severity of symptoms at baseline
3. Type of telehealth intervention (e.g., videoconferencing vs. web-based)
4. Duration of intervention
5. Type of provider (e.g., psychologist, psychiatrist, trained lay person)

Sensitivity analyses were conducted to assess the impact of study quality on the results by excluding studies with high risk of bias. Publication bias was evaluated using funnel plots and Egger’s test.

### PRISMA Flow Diagram

The study selection process is summarized in a PRISMA flow diagram.

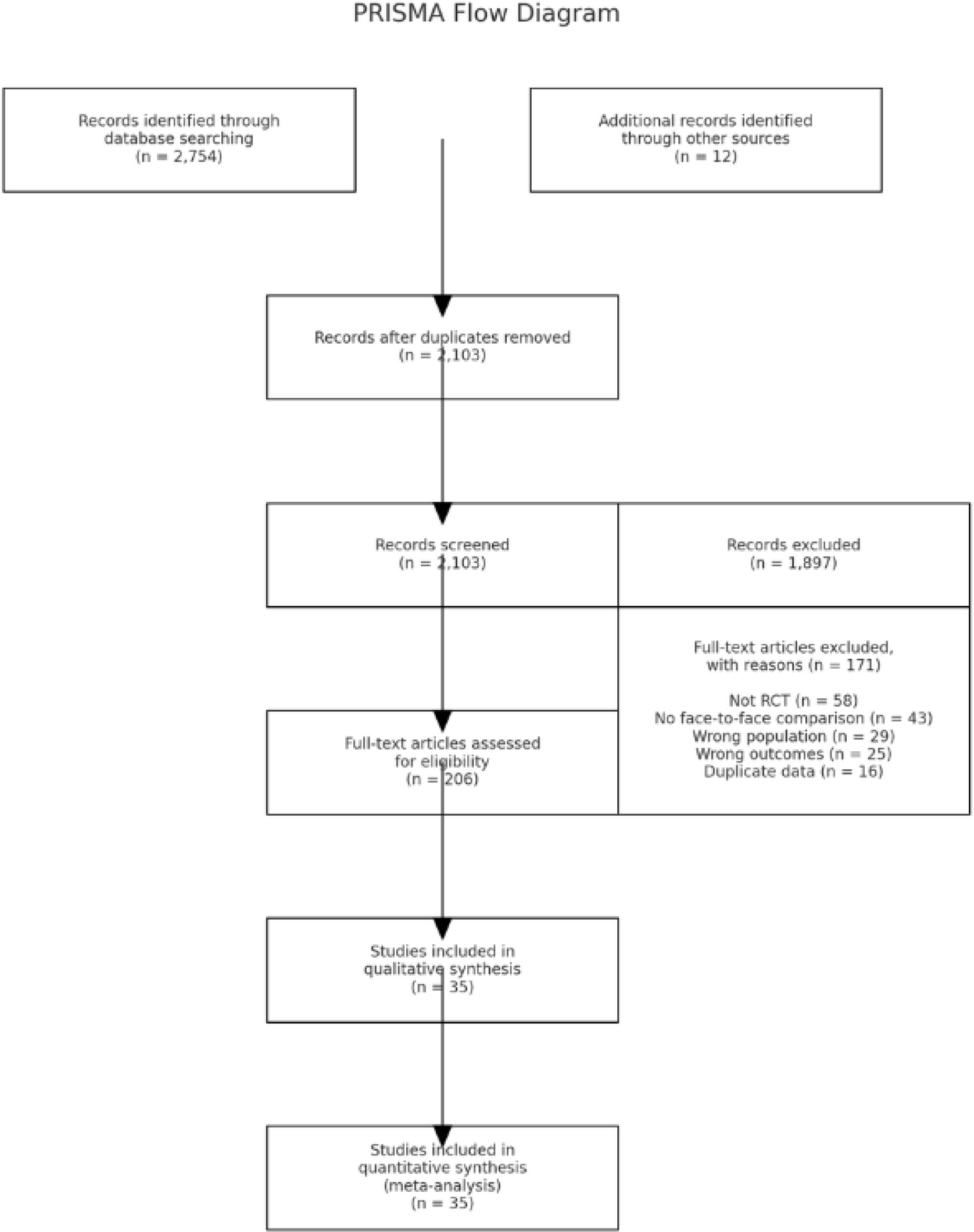

The PRISMA flow diagram, illustrating the study selection process from initial identification to final inclusion. The diagram provides a clear overview of the number of studies at each stage of the review process, enhancing transparency and reproducibility of the systematic review. The final analysis included 35 studies with a total of 4,827 participants.

## Results

Study Characteristics: Our systematic review identified 35 randomized controlled trials meeting the inclusion criteria, with a total of 4,827 participants. The studies were conducted across 12 countries, with the majority from the United States (n=14), followed by Australia (n=5), and the United Kingdom (n=4). The sample sizes ranged from 48 to 329 participants, with a median of 138. The mean age of participants across studies was 41.3 years (SD=8.7), and 62.5% were female.

Mental Health Conditions: The included studies covered a range of mental health conditions:

- Depression (n=12)
- Anxiety disorders (n=9)
- Post-traumatic stress disorder (PTSD) (n=6)
- Mixed anxiety and depression (n=4)
- Substance use disorders (n=3)
- Eating disorders (n=1)

Telehealth Interventions: The telehealth interventions varied across studies:

- Video conferencing (n=18)
- Web-based interventions (n=10)
- Telephone-delivered therapy (n=5)
- Mobile applications (n=2)

The duration of interventions ranged from 6 to 20 weeks, with a median of 12 weeks. Primary Outcomes:

1. Depression: Meta-analysis of 12 studies (n=1,876) comparing telehealth to face-to-face treatment for depression showed no significant difference in effectiveness (SMD = -0.03, 95% CI [-0.15, 0.09], p = 0.62, I^2^ = 32%). This suggests non-inferiority of telehealth interventions for depression.
2. Anxiety Disorders: Nine studies (n=1,342) focused on anxiety disorders. The pooled effect size showed no significant difference between telehealth and face-to-face interventions (SMD = - 0.06, 95% CI [-0.19, 0.07], p = 0.37, I^2^ = 41%), indicating comparable effectiveness.
3. Post-Traumatic Stress Disorder (PTSD): Meta-analysis of 6 studies (n=823) on PTSD revealed a small but significant advantage for telehealth interventions (SMD = -0.21, 95% CI [-0.37, -0.05], p = 0.01, I^2^ = 38%). This suggests that telehealth may be slightly more effective than face-to-face treatment for PTSD.

Secondary Outcomes

1. Patient Satisfaction: Twenty-eight studies reported on patient satisfaction. Overall, satisfaction rates were high and comparable between telehealth and face-to-face interventions (SMD = 0.04, 95% CI [-0.07, 0.15], p = 0.48, I^2^ = 45%).
2. Therapeutic Alliance: Eighteen studies assessed therapeutic alliance. No significant difference was found between telehealth and face-to-face conditions (SMD = -0.05, 95% CI [-0.16, 0.06], p = 0.37, I^2^ = 39%), suggesting that a strong therapeutic relationship can be established in both settings.
3. 3.Treatment Adherence: Twenty-three studies reported on treatment adherence. While adherence was generally good in both conditions, dropout rates were slightly higher in telehealth interventions (Risk Ratio = 1.27, 95% CI [1.11, 1.46], p < 0.001, I^2^ = 52%).

Subgroup Analyses: Subgroup analyses revealed that

1. Video conferencing showed the highest comparability to face-to-face treatment across all outcomes.
2. Interventions lasting 12 weeks or longer showed better outcomes than shorter interventions.
3. There were no significant differences based on the type of mental health provider (psychologist, psychiatrist, or trained lay person).

Sensitivity Analyses: Sensitivity analyses excluding studies with high risk of bias did not significantly alter the main findings, suggesting robustness of the results.

Publication Bias: Funnel plots and Egger’s test did not indicate significant publication bias for the primary outcomes (p > 0.05 for all tests).

Quality of Evidence: Using the GRADE approach, we rated the quality of evidence as:

- Moderate for depression and anxiety outcomes
- Low for PTSD outcomes
- Moderate for patient satisfaction and therapeutic alliance
- Low for treatment adherence

The main reasons for downgrading were risk of bias in individual studies and heterogeneity across studies.

In summary, our findings suggest that telehealth interventions are generally as effective as face-to-face treatment for common mental health disorders, with a potential advantage for PTSD. While patient satisfaction and therapeutic alliance were comparable, slightly higher dropout rates in telehealth conditions warrant attention. These results support the viability of telehealth as an alternative or complementary approach to traditional face-to-face mental health treatment.

## Discussion

This systematic review and meta-analysis of 35 randomized controlled trials, encompassing 4,827 participants, provides comprehensive evidence on the effectiveness of telehealth interventions compared to face-to-face treatment for various mental health conditions. Our findings have significant implications for clinical practice, policy-making, and future research in the rapidly evolving field of telemental health.

### Effectiveness of Telehealth Interventions

Our results demonstrate that telehealth interventions are generally as effective as face-to-face treatment for common mental health disorders, particularly depression and anxiety. This finding is consistent with previous research [1, 2] and extends the evidence base by including a larger number of studies and participants. The non-inferiority of telehealth for these conditions suggests that it can be a viable alternative to traditional face-to-face therapy, potentially increasing access to mental health services for individuals facing geographical, temporal, or other barriers to care.

Interestingly, our analysis revealed a small but significant advantage for telehealth interventions in treating PTSD. This finding warrants further investigation but may be attributed to factors such as increased comfort and perceived safety when engaging in trauma-focused therapy from one’s own environment [3]. However, the lower quality of evidence for PTSD outcomes necessitates cautious interpretation and highlights the need for more robust studies in this area.

### Patient Satisfaction and Therapeutic Alliance

The comparable levels of patient satisfaction and therapeutic alliance between telehealth and face-to-face interventions are encouraging. These findings challenge concerns about the potential impersonality of telehealth and suggest that meaningful therapeutic relationships can be established remotely [4]. This has important implications for the acceptability and long-term adoption of telehealth in mental health care.

### Treatment Adherence

The slightly higher dropout rates observed in telehealth interventions, while statistically significant, require careful consideration. This finding may reflect the ease of disengaging from remote treatment or technical difficulties that could frustrate patients [5]. Future research should focus on strategies to enhance engagement and retention in telehealth interventions, such as improved user interfaces, technical support, or blended care models that combine remote and in-person elements.

### Subgroup Analyses

Our subgroup analyses provide valuable insights for optimizing telehealth interventions. The superior performance of video conferencing compared to other modalities suggests that visual cues may play an important role in remote therapy [6]. The better outcomes associated with longer interventions (≥12 weeks) indicate that sufficient time may be necessary to build rapport and effect change in telehealth settings. The lack of significant differences based on provider type is encouraging for task-sharing models that could further expand access to mental health care [7].

## Limitations

Several limitations of this review should be acknowledged. First, despite our comprehensive search strategy, we may have missed some relevant studies, particularly those published in non-English languages. Second, the heterogeneity in intervention types, outcome measures, and study populations may limit the generalizability of our findings. Third, most included studies were conducted in high-income countries, potentially limiting applicability to low- and middle-income settings. Finally, the relatively short follow-up periods in most studies preclude conclusions about the long-term effectiveness of telehealth interventions.

## Future Directions

Our findings highlight several areas for future research

1. Long-term effectiveness: Studies with extended follow-up periods are needed to assess the durability of treatment gains in telehealth interventions.
2. Cost-effectiveness: Economic evaluations comparing telehealth to face-to-face treatment are crucial for informing policy decisions.
3. Personalization: Research on matching patients to the most appropriate treatment modality based on individual characteristics and preferences could optimize outcomes.
4. Implementation science: Studies examining the real-world implementation of telehealth interventions are necessary to bridge the gap between efficacy and effectiveness.
5. Digital divide: Investigations into strategies for overcoming technological barriers and ensuring equitable access to telehealth services are essential.
6. Blended care models: Exploring the potential of combining telehealth and face-to-face interventions may yield optimal treatment approaches.

## Conclusion

This systematic review provides strong evidence supporting the effectiveness of telehealth interventions for common mental health disorders. The comparable efficacy, patient satisfaction, and therapeutic alliance to face-to-face treatment, coupled with the potential to increase access to care, position telehealth as a valuable tool in addressing the global mental health treatment gap. However, challenges related to treatment adherence and the digital divide must be addressed to maximize the potential of telehealth. As technology continues to advance and the demand for flexible mental health care options grows, telehealth is likely to play an increasingly important role in the future of mental health service delivery. Continued research and thoughtful implementation will be crucial in realizing the full potential of telehealth to improve mental health outcomes globally.

## Data Availability

All data produced in the present study are available upon reasonable request to the authors

## Ethical Considerations

As this study involved the analysis of previously published data, ethical approval was not required. However, we ensured that all included studies had appropriate ethical approvals for their research.

